# Abortion Attitudes and Behavioral Intentions of Obstetrics and Gynecology Residents at Four Midwestern Residency Programs

**DOI:** 10.1101/2022.07.26.22278076

**Authors:** Abigail S. Cutler, Elise S. Cowley, Jessika A. Ralph, Jessie Chen, Amy Godecker, Jordan Ward, Sarah Hutto, Laura Jacques

## Abstract

In June 2022, the U.S. Supreme Court overturned *Roe v Wade*. Half of states now face proposed or in-effect abortion bans, which affect the ability of obstetrics and gynecology (ObGyn) residency programs to provide abortion training. Prior to the Supreme Court decision, we surveyed ObGyn residents at all four programs in Wisconsin and Minnesota to assess residents’ attitudes toward abortion, desire to learn about abortion, and intentions about providing abortion care in their future practice. We found that participants overwhelmingly support abortion, find the issue to be important, and plan to incorporate abortion into future practice. The reversal of *Roe v Wade* and its impact on access to abortion training may have implications for ObGyn residency recruitment and training, related career decision-making, and future workforce.

## Introduction

The reversal of *Roe V. Wade* by the United States Supreme Court on June 24, 2022 has resulted in proposed or in-effect abortion bans spanning half the country^1^. Despite being one of the most common medical procedures in the U.S. and a required component of Obstetrics and Gynecology (ObGyn) resident education by the Accreditation Council of Graduate Medical Education, nearly half of all ObGyn residency programs now struggle to provide clinical abortion training^2–5^. The media has also raised concerns that wide geographic variations in abortion legality will adversely shape where physicians choose to train and ultimately practice ^6–8^. To better understand how the fall of *Roe* may affect ObGyn residents’ career decisions, it is crucial to understand their current attitudes towards abortion, their desire to learn about abortion, and the importance they place on being able to provide abortion care in their future work. Current literature on these topics is sparse. We assessed attitudes and career intentions toward abortion among ObGyn residents in Minnesota and Wisconsin.

### Methods

We emailed a voluntary online survey to all ObGyn residents scheduled to participate in a workshop exploring abortion attitudes at the University of Minnesota (UMN), University of Wisconsin-Madison (UW), Medical College of Wisconsin (MCW) and Aurora-Sinai Milwaukee (Aurora) between January and December 2021. We obtained demographic information and used a previously published questionnaire to assess attitudes toward abortion care and behavioral intentions for future practice^9^. To assess attitudes, we asked the degree to which participants agreed with 17 statements about abortion using a 5-point Likert scale. To assess behavioral intentions, we posed six yes/no questions regarding intent to learn about, advocate for, refer patients to, and provide abortion care. Survey participants received a ten-dollar Amazon gift-card link. We created summative attitude scores which ranged from 0 (most negative toward abortion) to 100 (most positive toward abortion)^9^. We performed the Kruskal-Wallis rank test to assess for significant differences between observed institutional summative attitude scores.

## Results

A total of 55/70 (79%) ObGyn residents completed the survey, 17/21 (81%) from UMN, 14/20 (70%) from UW, 14/16 (88%) from MCW, and 10/13 (77%) from Aurora. Of residents who completed the survey, 46 (84%) identified as women, 28 (51%) were born in the Midwest, 36 (65%) did not identify with a particular religion (Table 1). Summaries of responses to individual attitude and behavioral intention questions are shown in Figure 1.

**Table 1.** Demographics of survey participants.

| **Resident characteristics** | **N (%)** |
| --- | --- |
| **Total** | **55 (100)** |
| **Institution** |  |
| University of Minnesota, Twin Cities | 17 (31) |
| Medical College of Wisconsin | 14 (25) |
| Aurora-Sinai of Milwaukee | 10 (18) |
| University of Wisconsin-Madison | 14 (25) |
| **Self-Identified Gender** |  |
| Man | 9 (16) |
| Woman | 46 (84) |
| **Age (years)** |  |
| Minimum | 26 |
| Maximum | 35 |
| Mean | 29 |
| **Born in the United States** |  |
| No | 5 (9) |
| Yes | 50 (91) |
| **Born in a Midwestern<sup>1</sup> State** |  |
| No | 22 (44) |
| Yes | 28 (56) |
| **Identify with a particular religion** |  |
| No | 36 (65) |
| Yes | 19 (35) |
| **Year in Training** |  |
| PGY1 | 17 (31) |
| PGY2 | 14 (25) |
| PGY3 | 12 (22) |
| PGY4 | 12 (22) |
| **Interested in pursuing a fellowship** |  |
| No | 39 (71) |
| Yes | 16 (29) |
| <sup>1</sup>Following U.S. Census definitions, Midwestern states included Illinois, Indiana, Iowa, Kansas, Michigan, Minnesota, Missouri, Nebraska, North Dakota, Ohio, South Dakota, and Wisconsin. |  |

**Figure 1.**
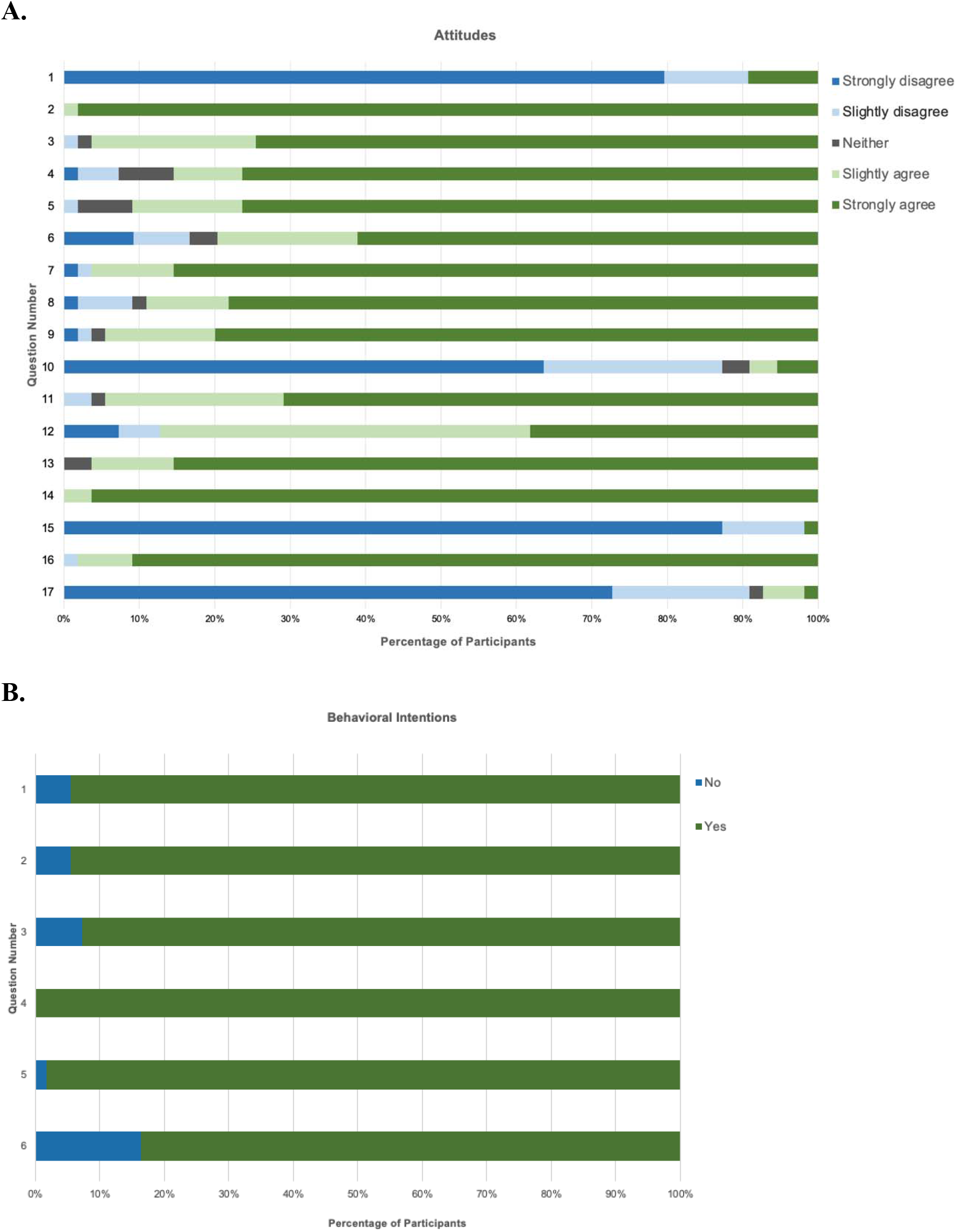
Summary of attitudes and behavioral intentions on abortion for all participants. Percentage of participants for each response option for **A**. attitudes and **B**. behavioral intentions on abortion.

The mean attitude score for ObGyn residents was 92.4 (SD 8.0). There were no differences in mean attitude scores among institutions. Nearly all residents (49/55, 89%) disagreed with the statement “the issue of abortion has little importance to me”. Almost all residents agreed that “All people should have access to safe, comprehensive abortion care in the first (55/55, 100%) and second (54/55, 98%) trimester. Nearly all residents (52/55, 95%) wanted to learn more about the need for safe, comprehensive abortion care and 46/55 (84%) planned to provide abortion care in their future careers.

## Discussion

Our research demonstrates that ObGyn residents in our sample hold highly favorable attitudes toward abortion, and nearly all find the issue of abortion to be important and plan to incorporate abortion care into their future work. This data suggests that access to abortion training may influence where ObGyn residents choose to train and ultimately practice. While some who value abortion training and provision may avoid states where abortion is restricted, others may wish to be drawn where the need for advocacy is high. Understanding the forces that shape the future ObGyn workforce is key, especially given pre-existing concerns about impending ObGyn shortages in certain areas of the country^10^. Future research should directly evaluate how state-level abortion restrictions impact both recruitment into ObGyn residency programs and career decision-making among graduating residents.

## Data Availability

All data produced in the present study are available upon reasonable request to the authors

## Acknowledgements

We wish to thank Andrea Zorbas, Sharon Blohowiak, Amanda Wildenberg, and Kelly Winum for administrative support. We thank Allison Linton and Kristina Kaljo for their contributions to study design and recruitment. We thank Nathan Jones and the UW-Madison Survey Center. The project was supported by institutional funding from the University of Wisconsin Department of Obstetrics and Gynecology. One author (ESC) was supported by a National Library training grant to the Computation and Informatics in Biology and Medicine Training Program (NLM 5T15LM007359) at UW-Madison and in part by Medical Scientist Training Program grant (T32GM140935). Some data from this study were previously presented in a poster at the 2022 CREOG & APGO Annual Meeting in Orlando, FL (March 2022).

## Attitude Questions

1. The issue of abortion is of little importance to me.
2. I support the provision of family planning and contraceptive services.
3. I feel comfortable working to increase access to family planning and contraceptive services.
4. I support the provision of abortion services as permitted by law.
5. I feel comfortable working to increase access to abortion services as permitted by law.
6. I feel comfortable talking with my closest family members about my involvement with abortion care.
7. I would feel comfortable observing an abortion procedure.
8. I would feel comfortable performing or assisting an abortion procedure.
9. I am clear about my personal values concerning abortion.
10. I feel very conflicted about abortion.
11. I can clearly explain my personal values concerning abortion.
12. I can respectfully explain values concerning abortion that conflict with mine.
13. I feel empathy for people who have experienced abortion.
14. All people should have access to safe, comprehensive abortion care in the first trimester.
15. Access to first trimester abortion should be restricted to certain circumstances.
16. All people should have access to safe, comprehensive abortion care in the second trimester.
17. Access to second trimester abortion should be restricted to certain circumstances.

## Behavioral Intentions Questions

1. Learn more about the need for safe, comprehensive abortion care.
2. Raise awareness about the need for safe, comprehensive abortion care.
3. Advocate making safe, comprehensive abortion care widely available.
4. Educate people about safe abortion services.
5. Refer people seeking abortion to safe services.
6. Provide or assist with safe, comprehensive abortion procedures.

